# Effects of Resveratrol as an Adjunct to a Low-Calorie Diet in Postmenopausal Women with Obesity and Knee Osteoarthritis

**DOI:** 10.64898/2026.06.09.26355282

**Authors:** Georgy Leonov, Anastasia Malvina, Svetlana Kosyura, Elena Livantsova, Yurgita Varaeva, Antonina Starodubova

**Author notes:** Correspondence: Georgy Leonov.

## Abstract

**Background:** Obesity is a modifiable risk factor for osteoarthritis and may contribute to pain, functional impairment, inflammation, and cartilage degradation. Resveratrol has potential anti-inflammatory and chondroprotective effects, but its efficacy as an adjunct to dietary intervention remains unclear.

**Objective:** This study evaluated whether resveratrol supplementation provides additional benefits when combined with a low-calorie diet in postmenopausal women with obesity and knee osteoarthritis.

**Methods:** A total of 97 postmenopausal women with obesity and knee osteoarthritis were included in this randomized controlled clinical study. Participants received either a 10-day low-calorie diet alone or the same diet combined with 150 mg/day trans-resveratrol. Anthropometric parameters, body composition, biochemical markers, pain intensity, functional status, and urinary CTX-II were assessed at baseline and follow-up.

**Results:** Both interventions were associated with reductions in body weight, BMI, waist and hip circumferences, fat mass, glucose, HOMA-IR, lipid parameters, hsCRP, VAS, WOMAC, LAI, and urinary CTX-II. Compared with diet alone, resveratrol supplementation did not provide additional benefits for anthropometric parameters, glucose metabolism, lipid profile, or WOMAC score. However, the resveratrol group showed a greater reduction in hsCRP and urinary CTX-II. The obesity class did not modify the treatment effect.

**Conclusion:** A short-term low-calorie diet improved metabolic, inflammatory, and osteoarthritis-related parameters in postmenopausal women with obesity and knee osteoarthritis. The addition of resveratrol did not enhance weight loss or improve most metabolic outcomes but was associated with greater reductions in hsCRP and urinary CTX-II. These findings suggest a potential anti-inflammatory and cartilage-related effect of resveratrol, which requires confirmation in longer randomized trials.

## Introduction

The ongoing global aging of the population has led to a growing concern regarding the burden of non-communicable chronic diseases (1). The prevalence of obesity has exhibited a consistent upward trend over the past three decades, and this condition now represents a significant contributing factor to rising healthcare costs. This phenomenon is accompanied by a concomitant decline in physical and mental well-being, as well as an increased susceptibility to the development of multiple chronic conditions (2). In the context of knee osteoarthritis (OA), obesity is a well-established modifiable risk factor that contributes to symptom severity (3,4). In addition, non-modifiable factors such as increasing age and female sex are consistently identified as major determinants of OA risk. The higher incidence of OA increases around the age of 50 years, which coincides with the onset of the menopausal transition. The increased prevalence of OA in postmenopausal women may be attributable to menopause-related hormonal changes, particularly declining estrogen levels (5,6). The knee is the most frequently affected weight-bearing joint, and knee OA is a significant cause of pain, disability, functional decline, and reduced quality of life (7).

The association between obesity and knee OA extends beyond mechanical overload. Obesity and metabolic syndrome have been associated with a state of chronic low-grade inflammation, increased expression of pro-inflammatory cytokines, altered adipokine secretion, up-regulation of proteolytic enzymes such as matrix metalloproteinases and aggrecanases, dyslipidemia-related increases in free fatty acids, and oxidative stress. These mechanisms may contribute to cartilage degradation and the progression of OA (8–11). Therefore, an effective treatment strategy for knee OA in patients with obesity should encompass the optimization of body weight and the consideration of metabolic and inflammatory pathways. Furthermore, many obese patients with severe osteoarthritis require joint replacement. Given the link between obesity and lower limb OA, as well as the need for joint replacement, clinical guidelines for OA management worldwide recommend weight management as a core strategy for all OA patients (12,13).

Resveratrol (3,4′,5-trihydroxystilbene) is a natural polyphenolic compound that is found in a variety of plants, including grape skins, grape seeds, Japanese knotweed, cassia seeds, and peanuts (14). Numerous studies have documented its potential to exert anti-inflammatory, antioxidant, antiplatelet, antihyperlipidemic, cardioprotective, vasorelaxant, neuroprotective, and estrogen-like effects (15–17). A substantial body of experimental and clinical research suggests that resveratrol may possess therapeutic potential in OA through several mechanisms, including the protection of chondrocytes, the regulation of inflammatory signaling, the inhibition of chondrocyte apoptosis, the preservation of the subchondral bone microenvironment, and the reduction of extracellular matrix degradation (18–20). Although weight management is recommended as a core component of knee OA care, the short-term effects of calorie restriction on pain, function, and cartilage degradation biomarkers remain insufficiently characterized in postmenopausal women with severe obesity. Moreover, it remains unclear whether resveratrol can provide additional benefit beyond dietary restriction in this high-risk phenotype (21).

In this controlled clinical trial, we investigated whether resveratrol supplementation provides additional benefit when combined with a low-calorie diet in postmenopausal women with obesity and knee osteoarthritis.

## Methods

### Study design

This controlled clinical trial evaluated the effect of resveratrol supplementation combined with a low-calorie diet in postmenopausal women with obesity and knee osteoarthritis. Study subjects were assigned to either a low-calorie diet group or a low-calorie diet plus resveratrol supplementation group. Participants were randomized into two groups in a 1:1 ratio using a simple randomization procedure. The allocation sequence was generated before the intervention using Research Randomizer software (https://www.randomizer.org/). Allocation concealment was maintained with sequentially numbered, opaque, sealed envelopes opened after eligibility confirmation and baseline assessment.

The primary outcomes included changes in pain, functional status, systemic inflammation, and urinary CTX-II. Secondary outcomes included anthropometric parameters, body composition, biochemical and metabolic markers. Assessments were performed at baseline and follow-up and included anthropometry, body composition analysis, blood biochemical testing, Visual Analog Scale (VAS), Western Ontario and McMaster Universities Osteoarthritis Index (WOMAC), Lequesne Algofunctional Index (LAI), and urinary C-terminal telopeptide of type II collagen (urinary CTX-II). The study was retrospectively registered at ClinicalTrials.gov under the identifier NCT07634302.

### Study participants

Consecutive eligible participants were recruited during inpatient treatment at the Nutrition Clinic of the Federal Research Centre of Nutrition, Biotechnology and Food Safety. All participants were postmenopausal women aged 45–75 years with obesity, defined as BMI ≥30 kg/m². Knee osteoarthritis was diagnosed according to standard clinical and radiographic criteria, including knee pain and radiographic signs of osteoarthritis. The study included only participants with Kellgren-Lawrence grade 2 or 3 knee osteoarthritis. Postmenopausal status was defined as the absence of menstruation for at least 12 consecutive months.

Participants were assigned to the low-calorie diet group (control group, n = 52) or the low-calorie diet plus resveratrol group (resveratrol group, n = 45). The mean age was 60.6 ± 6.87 years, and the mean BMI was 43.6 ± 7.05 kg/m². All participants completed baseline and follow-up assessments. Concomitant medications were continued during the study when clinically indicated. No new weight-loss medications, intra-articular injections, or chondroprotective agents were prescribed during the intervention. Inclusion and exclusion criteria, as well as group stratification, are shown in Figure 1.

**Figure 1.**
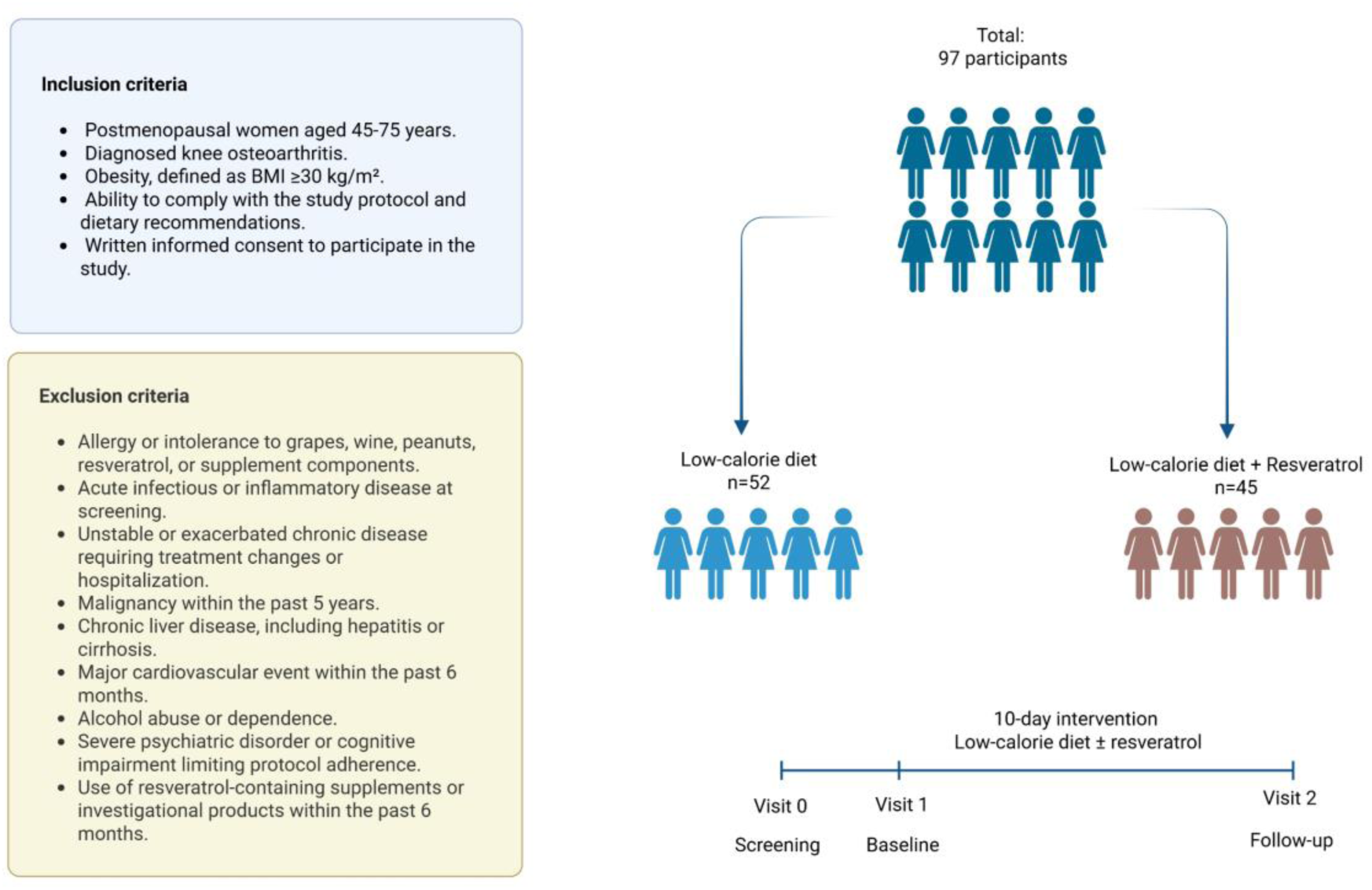
Flowchart of the study design and participant allocation. A total of 97 postmenopausal women with obesity and knee osteoarthritis were included in the study and allocated to either a low-calorie diet group or a low-calorie diet plus resveratrol group. Inclusion and exclusion criteria are shown.

The study was conducted in accordance with the Declaration of Helsinki and was approved by the Local Ethics Committee of the Federal Research Centre of Nutrition, Biotechnology and Food Safety (protocol code N1/2017 dated on 03/FEB/2017). All participants provided written informed consent before enrollment.

### Anthropometry, body composition and biochemical indicators

Anthropometric and body composition assessments were performed at baseline and follow-up. Body weight and height were measured using standard procedures, and body mass index was calculated as weight in kilograms divided by height in meters squared. Waist and hip circumferences were also measured.

Body composition was assessed by multifrequency bioelectrical impedance analysis using ABC-01 Medass (Medass Ltd, Russia). The assessed parameters included basal metabolic rate, fat mass, fat percentage and lean mass.

Blood samples were collected at baseline (BL) and follow-up (FU). Serum total cholesterol (TC), low-density lipoprotein cholesterol (LDL-C), high-density lipoprotein cholesterol (HDL-C), triglycerides (TG), alanine aminotransferase (ALT), aspartate aminotransferase (AST), urea, creatinine, uric acid, glucose, insulin, and high-sensitivity C-reactive protein (hsCRP) were measured in the clinical biochemistry laboratory using a KONELAB Prime 60i analyzer (Thermo Fisher Scientific, Waltham, MA, USA). HOMA-IR was calculated using fasting glucose and insulin concentrations. In addition, the TG to HDL-C ratio was calculated (22).

Pain intensity was assessed using a 10-point visual analog scale, with higher scores indicating more severe pain. Functional status and osteoarthritis severity were evaluated using the WOMAC and the LAI (23,24).

Urinary CTX-II was measured as a marker of type II collagen degradation. Urine samples were collected before and after the intervention, and CTX-II concentrations were determined using the Urine CartiLaps ELISA kit (IDS Ltd., USA). Results were corrected for urinary creatinine concentration.

### Diet and supplementation protocol

During inpatient treatment, all participants received a 10-day low-calorie diet. The average energy value was ∼1700 kcal/day; protein 90 g/day (21.1%), fat 72 g/day (38.1%), and carbohydrates 173 g/day (40.8%). The diet was based on moderate energy restriction, mainly through reduced fat and carbohydrate intake. Added sugars were excluded, animal fats and sodium were restricted, and dietary fiber sources, including vegetables, fruits, and cereals, were included. Meals were prepared boiled or steamed, without added salt, and provided in a fractional meal. The low-calorie diet was designed to provide an energy intake below the estimated basal metabolic rate, taking into account the additional energy expenditure for the physical activity category. Dietary adherence was monitored by clinical staff because all meals were provided during inpatient treatment.

Participants in the control group received the low-calorie diet alone. Participants in the resveratrol group received the same low-calorie diet plus 150 mg/day >99% trans-resveratrol powder (DSM, Netherlands), administered once daily with a meal throughout the 10-day intervention.

### Statistical analysis

Statistical analyses were performed using IBM SPSS Statistics v22 (IBM Corp., Armonk, NY, USA) and GraphPad Prism v9 (San Diego, CA, USA). Continuous variables are presented as mean ± standard deviation, and categorical variables as n (%). Between-group differences at baseline were assessed using independent-samples t-tests or chi-square tests, as appropriate.

Within-group changes from baseline to follow-up were analyzed using paired-samples t-tests. Change values were calculated as follow-up minus baseline and are reported with 95% confidence intervals. Between-group differences at follow-up were evaluated using ANCOVA, with treatment group as the fixed factor and the corresponding baseline value as a covariate. Adjusted means, 95% confidence intervals, p values, and partial eta squared were reported.

Exploratory subgroup analyses were performed by obesity class using ANCOVA models including treatment group, obesity class, their interaction, and the corresponding baseline value as a covariate. Exploratory Spearman correlation analysis was used to assess associations between changes in selected outcomes. Standardized adjusted between-group differences were calculated for graphical presentation by dividing adjusted mean differences by the pooled baseline standard deviation. All tests were two-sided, and p < 0.05 was considered statistically significant.

## Results

### Participant characteristics

The study involved a total of 97 Caucasian postmenopausal women with obesity and knee osteoarthritis. The prevalence of hypertension was found to be 94 (96.9%), coronary artery disease – 14 (14.4%), dyslipidemia – 45 (46.4%), type 2 diabetes mellitus (T2DM) – 25 (25.8%), and non-alcoholic fatty liver disease (NAFLD) – 41 (42.3%). No statistically significant differences were observed between the groups (p > 0.05). As indicated by the findings of obesity classes, the most prevalent category was class III (BMI ≥ 40.0). The findings of the comparative analysis by group are presented in Table 1.

**Table 1.**
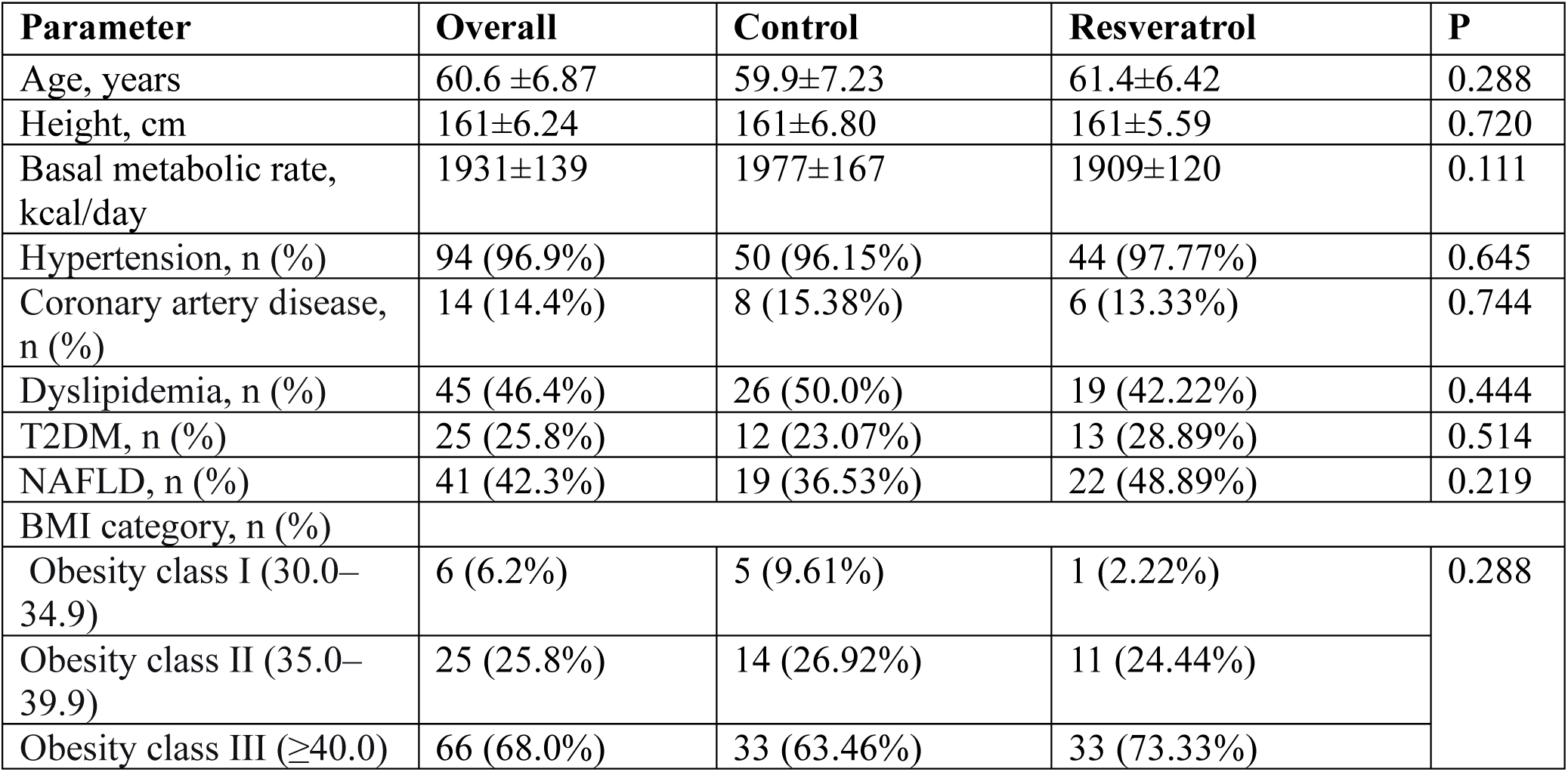
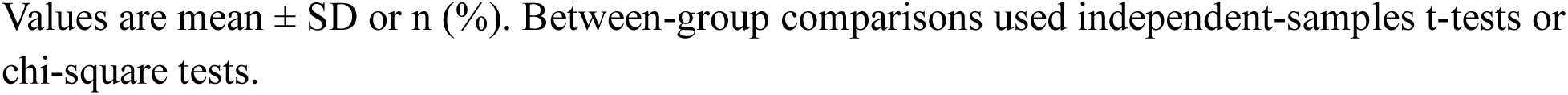
Baseline characteristics of the study participants.

| Parameter | Overall | Control | Resveratrol | P |
| --- | --- | --- | --- | --- |
| Age, years | 60.6 ± 6.87 | 59.9 ± 7.23 | 61.4 ± 6.42 | 0.288 |
| Height, cm | 161 ± 6.24 | 161 ± 6.80 | 161 ± 5.59 | 0.720 |
| Basal metabolic rate, kcal/day | 1931 ± 139 | 1977 ± 167 | 1909 ± 120 | 0.111 |
| Hypertension, n (%) | 94 (96.9%) | 50 (96.15%) | 44 (97.77%) | 0.645 |
| Coronary artery disease, n (%) | 14 (14.4%) | 8 (15.38%) | 6 (13.33%) | 0.744 |
| Dyslipidemia, n (%) | 45 (46.4%) | 26 (50.0%) | 19 (42.22%) | 0.444 |
| T2DM, n (%) | 25 (25.8%) | 12 (23.07%) | 13 (28.89%) | 0.514 |
| NAFLD, n (%) | 41 (42.3%) | 19 (36.53%) | 22 (48.89%) | 0.219 |
| BMI category, n (%) |  |  |  |  |
| Obesity class I (30.0–34.9) | 6 (6.2%) | 5 (9.61%) | 1 (2.22%) | 0.288 |
| Obesity class II (35.0–39.9) | 25 (25.8%) | 14 (26.92%) | 11 (24.44%) |  |
| Obesity class III ($\geq 40.0$) | 66 (68.0%) | 33 (63.46%) | 33 (73.33%) | |

Values are mean ± SD or n (%). Between-group comparisons used independent-samples t-tests or chi-square tests.

### Clinical outcomes

In addition, the groups did not differ in weight (p=0.126) and BMI (p=0.209), fat mass, serum glucose level, and lipid metabolism parameters (p>0.05). However, differences were found in waist circumference (p=0.036), hip circumference (p=0.022), and HOMA-IR (p=0.001). Adherence to a low-calorie diet (control group) and a low-calorie diet in combination with resveratrol (Resveratrol group) for 10 days led to significant changes in both anthropometric parameters and biochemical markers (Table 2). The investigation revealed a decline in body weight, BMI, waist and hip circumferences, fat mass, lean mass, uric acid, glucose, total cholesterol, triglycerides, non-HDL cholesterol, HOMA-IR, and hsCRP in both the control and resveratrol groups (all p = 0.001, except uric acid in the resveratrol group, p = 0.048, and glucose in the resveratrol group, p = 0.011). An increase in fat percentage was observed in the resveratrol group (p = 0.007), whereas the control group showed no statistically significant change (p = 0.212). The study found that the lean mass percentage remained unchanged in both groups (control, p = 0.502; resveratrol, p = 0.060). AST and ALT levels exhibited a decrease exclusively within the control group (AST, p = 0.001; ALT, p = 0.003), while no alterations were discerned in the resveratrol group (AST, p = 0.563; ALT, p = 0.941). However, both pre- and post-therapy parameters remained within normal limits. Creatinine levels demonstrated stability across both groups (control, p = 0.830; resveratrol, p = 0.051). HDL-C decreased in the resveratrol group (p = 0.001), with no statistically confirmed change in the control group (p = 0.052). The TG/HDL ratio remained unchanged in both groups (control, p = 0.331; resveratrol, p = 0.276).

**Table 2.** Changes in anthropometric, body composition, biochemical, and metabolic parameters.

| Parameter | Group | Baseline | Follow-up | $\Delta$ (95% CI) | P |
| --- | --- | --- | --- | --- | --- |
| Weight, kg | Control | 110.38±18.82 | 106.19±17.57 | -4.19 (-4.82; -3.55) | **0.001** |
| | Resveratrol | 115.88±15.85 | 111.92±15.11 | -3.97 (-4.47; -3.45) | **0.001** |
| P |  | 0.126 |  |  |  |
| BMI, kg/m<sup>2</sup> | Control | 42.79±7.71 | 41.16±7.21 | -1.59 (-1.71; -1.32) | **0.001** |
| | Resveratrol | 44.53±5.46 | 43.01±5.22 | -1.48 (-1.69; -1.28) | **0.001** |
| P |  | 0.209 |  |  |  |
| WC, cm | Control | 115.08±14.27 | 111.58±13.71 | -3.50 (-4.09; -2.90) | **0.001** |
| | Resveratrol | 120.80±11.92 | 116.53±11.13 | -4.26 (-5.16; -3.36) | **0.001** |
| P |  | **0.036** |  |  |  |
| HC, cm | Control | 125.23±15.71 | 121.83±14.51 | -3.40 (-4.23; -2.57) | **0.001** |
|  | Resveratrol | 131.76±10.93 | 128.40±10.33 | -3.35 (-4.01; -2.66) | **0.001** |
| P |  | **0.022** |  |  |  |
| Fat, % | Control | 49.23±5.69 | 49.45±5.82 | 0.22 (-0.13; 0.57) | 0.212 |
|  | Resveratrol | 49.70±3.78 | 50.34±3.59 | 0.64 (0.18; 1.11) | **0.007** |
| P |  | 0.642 |  |  |  |
| Fat, kg | Control | 54.53±12.66 | 52.75±12.30 | -1.78 (-2.26; -1.30) | **0.001** |
|  | Resveratrol | 58.01±11.22 | 56.74±10.82 | -1.26 (-1.76; -0.76) | **0.001** |
| P |  | 0.160 |  |  |  |
| Lean mass, % | Control | 52.09±5.32 | 52.30±5.50 | 0.20 (-0.41; 0.83) | 0.502 |
|  | Resveratrol | 50.31±3.64 | 49.81±3.62 | -0.50 (-1.03; 0.02) | 0.060 |
| P |  | 0.062 |  |  |  |
| Lean mass, kg | Control | 56.94±8.62 | 54.89±7.23 | -2.05 (-2.88; -1.22) | **0.001** |
|  | Resveratrol | 57.90±5.81 | 55.36±5.41 | -2.54 (-3.21; -1.87) | **0.001** |
| P |  | 0.531 |  |  |  |
| AST, U/L | Control | 22.29±10.30 | 19.28±9.92 | -3.01 (-4.11; -1.90) | **0.001** |
|  | Resveratrol | 21.61±6.88 | 21.09±7.62 | -0.52 (-2.34; 1.29) | 0.563 |
| P |  | 0.708 |  |  |  |
| ALT, U/L | Control | 22.79±13.62 | 20.64±12.03 | -2.14 (-3.56; -0.76) | **0.003** |
|  | Resveratrol | 21.03±8.06 | 21.11±10.79 | 0.08 (-2.18; 2.35) | 0.941 |
| P |  | 0.450 |  |  |  |
| Creatinine, μmol/L | Control | 63.67±12.83 | 64.02±10.46 | 0.34 (-2.87; 3.56) | 0.830 |
|  | Resveratrol | 66.03±14.62 | 63.84±11.11 | -2.18 (-4.37; 0.01) | 0.051 |
| P |  | 0.401 |  |  |  |
| Uric acid, μmol/L | Control | 359.58±80.94 | 337.60±75.95 | -21.98 (-25.9; -18.1) | **0.001** |
|  | Resveratrol | 352.20±96.75 | 338.31±95.33 | -13.88 (-27.6; -0.10) | **0.048** |
| P |  | 0.684 |  |  |  |
| Glucose, mmol/L | Control | 6.17±1.31 | 5.62±1.04 | -0.54 (-0.73; -0.36) | **0.001** |
|  | Resveratrol | 6.03±1.25 | 5.70±1.05 | -0.33 (-0.58; -0.08) | **0.011** |
| P |  | 0.604 |  |  |  |
| TC, mmol/L | Control | 5.42±1.05 | 5.00±1.02 | -0.41 (-0.56; -0.26) | **0.001** |
|  | Resveratrol | 5.42±1.09 | 4.91±0.82 | -0.50 (-0.70; -0.29) | **0.001** |
| P |  | 0.998 |  |  |  |
| TG, mmol/L | Control | 1.81±0.82 | 1.63±0.74 | -0.18 (-0.25; -0.11) | **0.001** |
|  | Resveratrol | 1.66±0.77 | 1.42±0.79 | -0.23 (-0.32; -0.15) | **0.001** |
| P |  | 0.362 |  |  |  |
| LDL-C, mmol/L | Control | 3.90±0.86 | 3.71±1.63 | -0.19 (-0.61; 0.23) | 0.371 |
|  | Resveratrol | 3.70±0.93 | 3.40±0.79 | -0.29 (-0.45; -0.13) | **0.001** |
| P |  | 0.288 |  |  |  |
| HDL-C, | Control | 1.13±0.29 | 1.05±0.28 | -0.08 (-0.16; 0.00) | 0.052 |
| mmol/L | Resveratrol | 1.17±0.25 | 1.05±0.17 | -0.12 (-0.17; -0.06) | **0.001** |
| P |  | 0.475 |  |  |  |
| Non-HDL<br>mmol/L | Control | 4.28±0.99 | 3.95±0.98 | -0.33 (-0.45; -0.19) | **0.001** |
|  | Resveratrol | 4.24±1.00 | 3.861±0.79 | -0.38 (-0.57; -0.18) | **0.001** |
| P |  | 0.839 |  |  |  |
| TG/HDL ratio | Control | 1.69±0.76 | 1.63±0.68 | -0.06 (-0.18; 0.06) | 0.331 |
|  | Resveratrol | 1.49±0.75 | 1.41±0.85 | -0.07 (-0.21; 0.06) | 0.276 |
| P |  | 0.207 |  |  |  |
| HOMA-IR | Control | 3.82±1.91 | 3.22±1.31 | -0.60 (-0.83; -0.36) | **0.001** |
|  | Resveratrol | 5.88±3.25 | 4.65±2.33 | -1.23 (-1.71; -0.74) | **0.001** |
| P |  | **0.001** |  |  |  |
| hsCRP, mg/L | Control | 5.15±2.82 | 4.70±2.68 | -0.44 (-0.60; -0.28) | **0.001** |
|  | Resveratrol | 5.66±3.95 | 4.76±3.81 | -0.90 (-1.23; -0.57) | **0.001** |
| P |  | 0.459 |  |  |  |

Values are mean ± SD. Between-group comparisons used independent-samples t-tests. Within-group changes were assessed using paired-samples t-tests.

The baseline indicators and markers of osteoarthritis were assessed in the study participants (Table 3). The study groups were found to differ in their functional joint state, as measured by WOMAC. Furthermore, a higher level of CTX-II in the urine was observed in the Resveratrol group. Following the conclusion of the treatment period, a significant decrease in severity indices of osteoarthritis (VAS, WOMAC, LAI), as well as levels of urinary CTX-II was observed in both groups (p<0.05).

**Table 3.** Changes in pain, functional outcomes, and urinary CTX-II.

| Parameter | Group | Baseline | Follow-up | Δ (95% CI) | P |
| --- | --- | --- | --- | --- | --- |
| VAS | Control | 6.96±1.26 | 5.75±1.10 | -1.21 (-1.42; -1.00) | **0.001** |
|  | Resveratrol | 7.04±1.31 | 6.11±1.32 | -0.93 (-1.16; -0.70) | **0.001** |
| P |  | 0.753 |  |  |  |
| WOMAC | Control | 176.52±21.29 | 161.50±22.32 | -15.01 (-17.28; -12.75) | **0.001** |
|  | Resveratrol | 190.07±22.87 | 174.07±23.26 | -16.00 (-19.41; -12.58) | **0.001** |
| P |  | **0.003** |  |  |  |
| LAI | Control | 10.08±1.91 | 8.73±1.86 | -1.34 (-1.52; -1.16) | **0.001** |
|  | Resveratrol | 10.29±1.64 | 8.96±1.83 | -1.33 (-1.59; -1.07) | **0.001** |
| P |  | 0.564 |  |  |  |
| CTX-II,<br>ng/mmol Cr | Control | 511.61±197.51 | 423.50±197.75 | -88.10 (-111.56; -64.65) | **0.001** |
|  | Resveratrol | 728.97±156.99 | 583.89±189.87 | -145.07 (-177.49; -112.66) | **0.001** |
| P |  | **0.001** |  |  |  |

Values are mean ± SD. Within-group changes were assessed using paired-samples t-tests

We conducted a comparative analysis of the efficacy of resveratrol supplementation in conjunction with a low-calorie diet as a standalone intervention (Table 4). In order to reduce the influence of baseline parameters on the results, an ANCOVA analysis was performed with baseline parameters as a covariate. Patients in the Resveratrol group experienced a slightly greater reduction in percentage lean mass (p=0.017). Conversely, a decline in blood AST levels was exclusively observed in the Control group (p=0.019), with levels remaining within the normal range. A more pronounced reduction in hsCRP was noted after 10 days of treatment in the Resveratrol group (p=0.013).

**Table 4.**
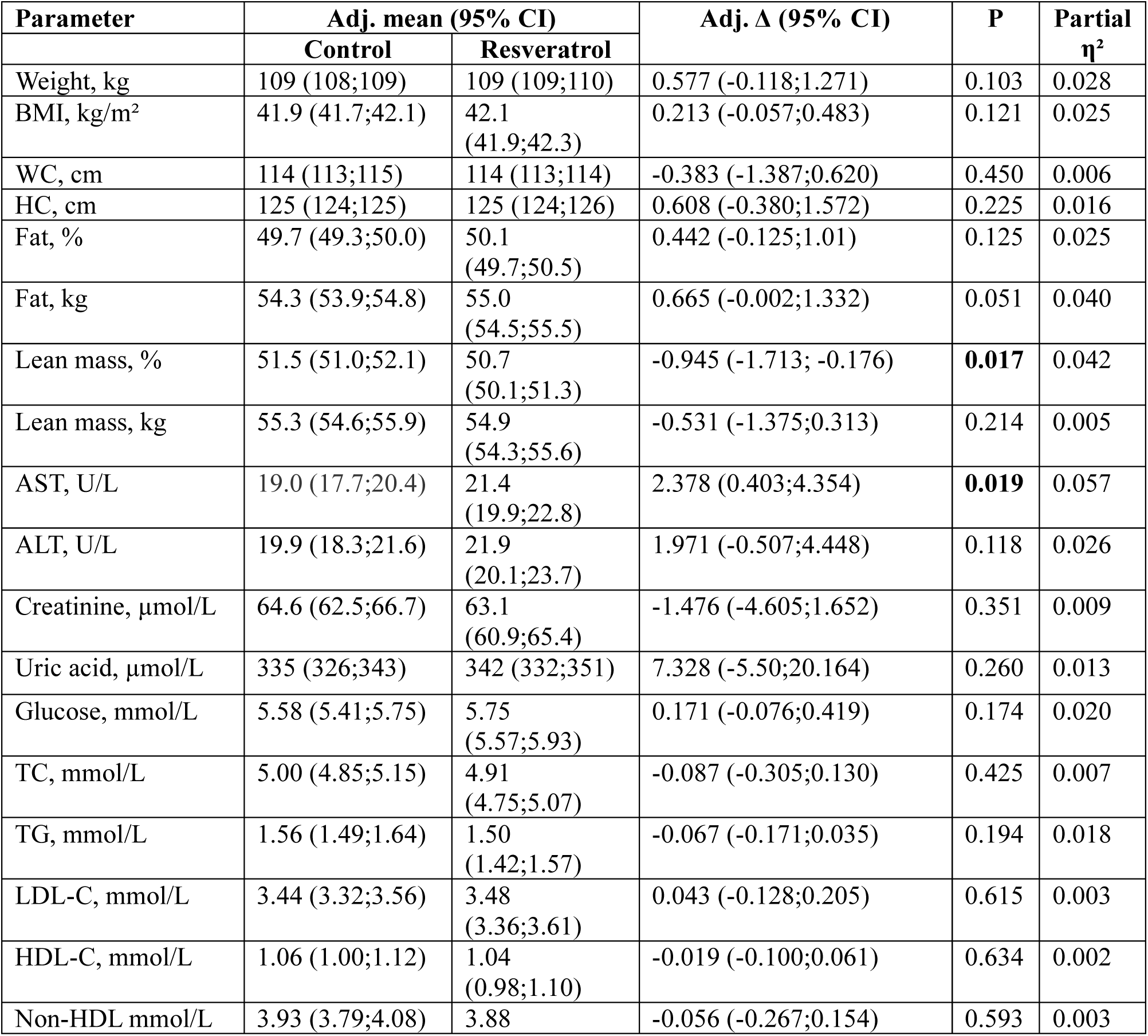
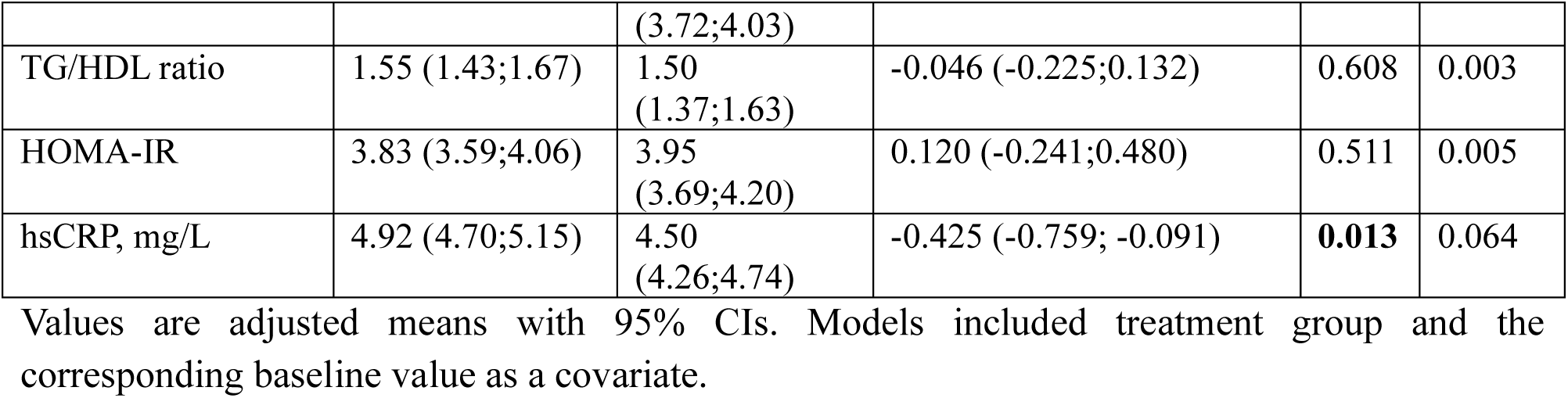
ANCOVA-adjusted follow-up values for anthropometric, body composition, biochemical, and metabolic parameters.

The analysis revealed that the changes in the WOMAC Osteoarthritis Functional Score were not significantly different between the groups (p=0.960). The reduction in pain score (VAS) was greater in the Control group (p=0.038). Meanwhile, urinary CTX-II levels decreased more significantly in the Resveratrol group (p=0.048). The data are presented in Figure 2.

**Figure 2.**
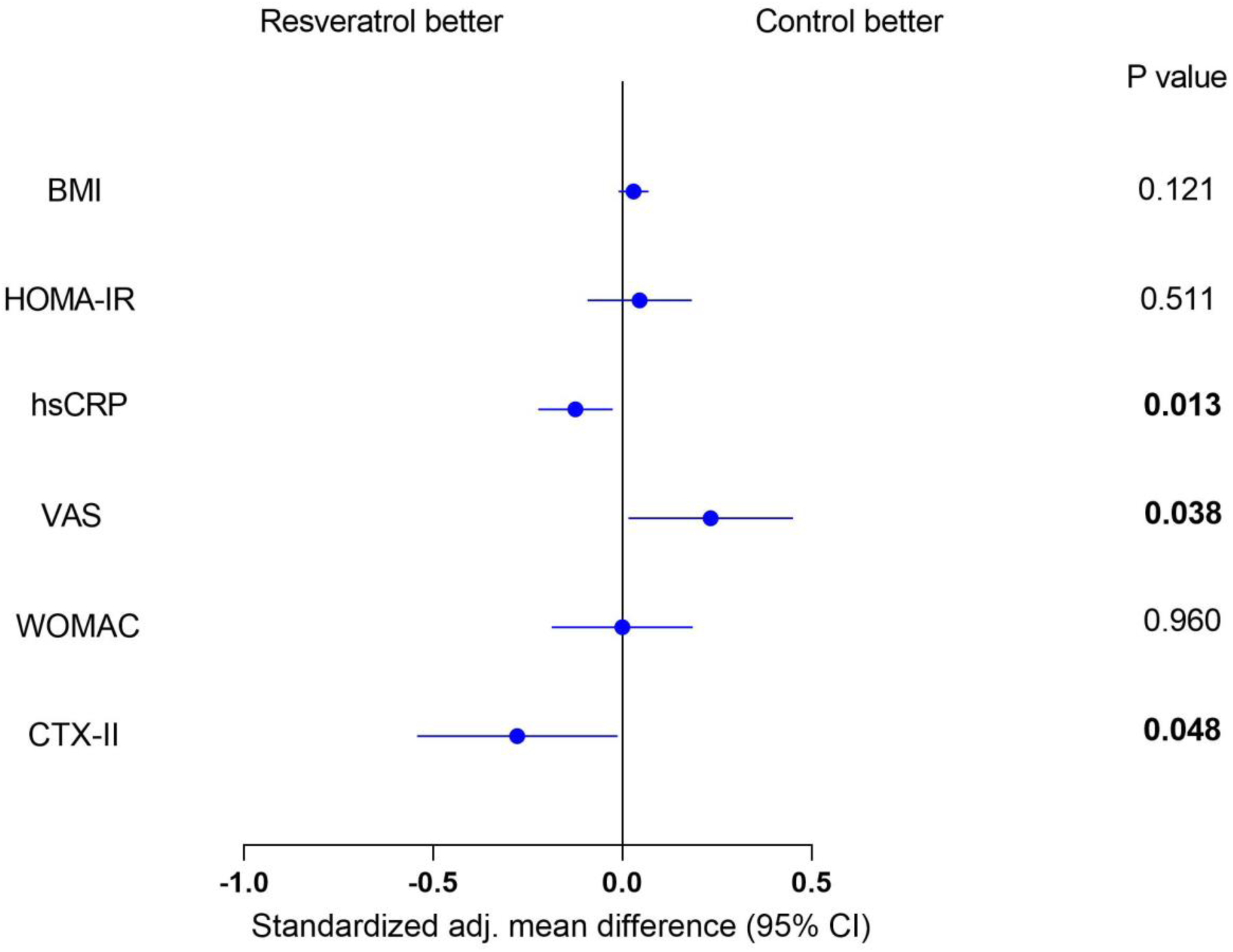
ANCOVA-adjusted standardized differences between the resveratrol and control groups.

To ascertain whether obesity severity influenced the study results, a subgroup analysis was conducted (Table 5). Participants were divided into two groups based on their obesity level: moderate (classes I and II) and severe (class III). The present study found no evidence that the severity of obesity influenced the efficacy of resveratrol (p>0.05).

**Table 5.** Exploratory ANCOVA-based subgroup analysis by obesity class.

| Parameter | Obesity subgroup | Adj. mean (95% CI) | | P | Partial $\eta^2$ |
| --- | --- | --- | --- | --- | --- |
|  |  | Control | Resveratrol |  |  |
| Weight, kg | Class I & II | 108 (107;109) | 109 (108;110) | 0.955 | 0.000 |
|  | Class III | 109 (108;109) | 109 (109;110) |  |  |
| WC, cm | Class I & II | 114 (112;115) | 113 (113;115) | 0.951 | 0.000 |
|  | Class III | 114 (113;115) | 114 (113;115) |  |  |
| Glucose, mmol/L | Class I & II | 5.50 (5.22;5.78) | 5.67 (5.32;6.02) | 0.939 | 0.000 |
|  | Class III | 5.62 (5.41;5.84) | 5.78 (5.56;5.99) |  |  |
| TC, mmol/L | Class I & II | 4.91 (4.66;5.15) | 4.84 (4.52;5.15) | 0.846 | 0.000 |
|  | Class III | 5.05 (4.87;5.24) | 4.94 (4.75;5.13) |  |  |
| HOMA-IR | Class I & II | 3.72 (3.32;4.11) | 3.90 (3.40;4.40) | 0.781 | 0.001 |
|  | Class III | 3.89 (3.60;4.18) | 3.97 (3.67;4.26) |  |  |
| hsCRP, mg/L | Class I & II | 4.85 (4.47;5.23) | 4.40 (3.92;4.88) | 0.959 | 0.000 |
|  | Class III | 4.97 (4.68;5.25) | 4.54 (4.25;4.82) |  |  |
| VAS | Class I & II | 5.70 (5.38;6.01) | 5.88 (5.48;6.28) | 0.639 | 0.002 |
|  | Class III | 5.83 (5.59;6.07) | 6.15 (5.91;6.39) |  |  |
| WOMAC | Class I & II | 169 (164;173) | 166 (161;172) | 0.365 | 0.009 |
|  | Class III | 167 (163;170) | 168 (164;171) |  |  |
| CTX-II, ng/mmol Cr | Class I & II | 558 (501;616) | 498 (490;569) | 0.713 | 0.002 |
|  | Class III | 530 (490;569) | 486 (452;520) |  |  |

Values are adjusted means with 95% CIs. Models included treatment group, obesity class, their interaction, and the corresponding baseline value as a covariate.

Furthermore, in order to evaluate the association between alterations in anthropometric and metabolic parameters and a reduction in osteoarthritis markers, a correlation analysis was conducted (Figure 3). Elevated serum HDL-C levels among participants were found to be associated with decreased urinary CTX-II levels (p<0.05).

**Figure 3.**
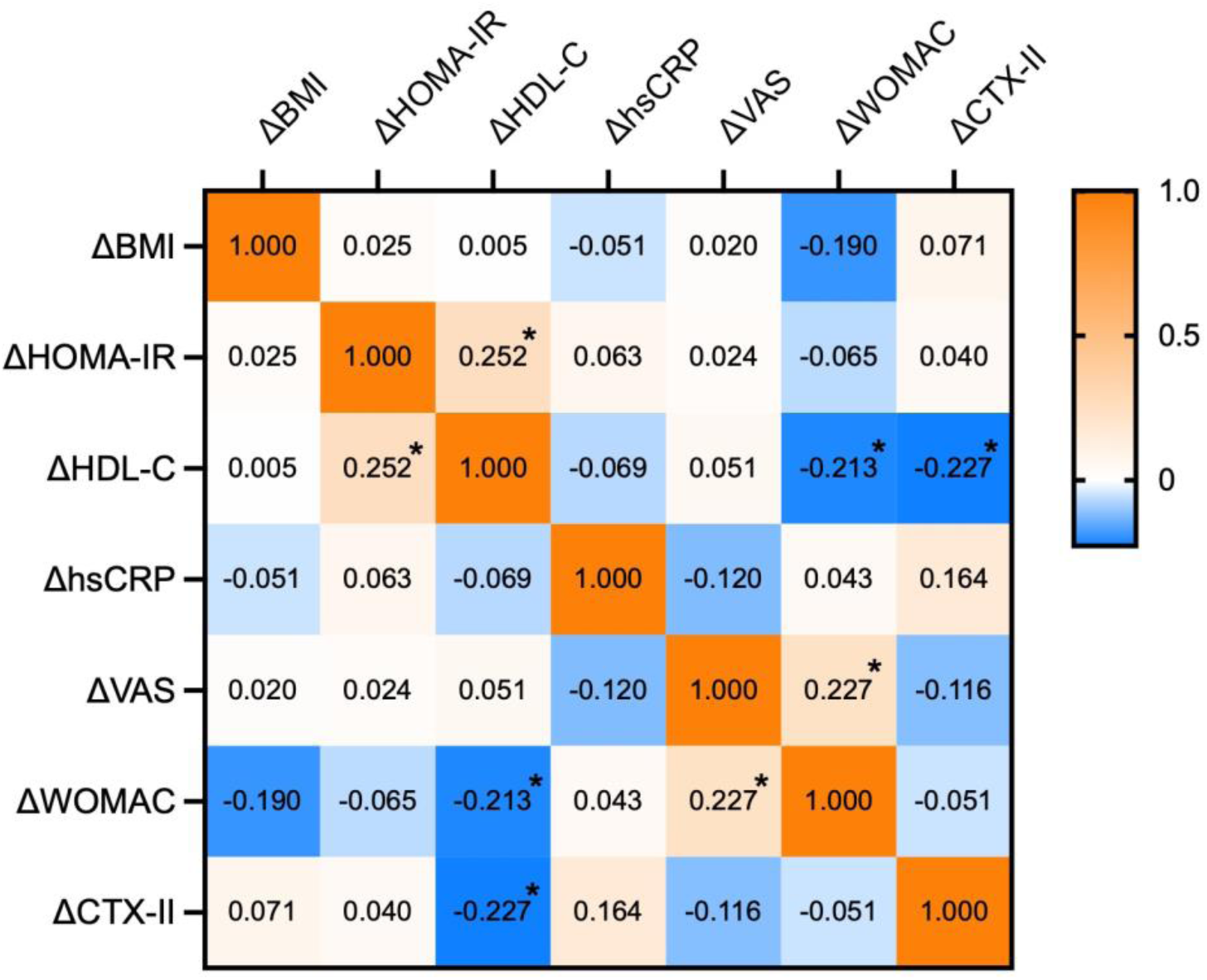
Exploratory Spearman correlation heatmap of changes in selected outcomes. * p ≤ 0.05.

## Discussion

In this study, it was demonstrated that adherence to a low-calorie diet (∼ 1700 kcal/day) over 10 days resulted in significant improvements in anthropometric parameters (weight, BMI, WC, and HC). However, it is notable that many patients also experienced a decrease in lean body mass. Furthermore, improvements in blood biochemical parameters, including markers of carbohydrate and lipid metabolism, were also observed. Specifically, the low-calorie diet resulted in a reduction of glucose levels, the HOMA-IR index, and TC, TG, LDL-C, HDL-C, and non-HDL levels. Concurrently, levels of the inflammatory marker hsCRP also decreased. Furthermore, parameters associated with the progression of knee osteoarthritis also exhibited significant improvement following a 10-day treatment period. Participants noted a reduction in pain, improvements in the functional assessment of the joint, and a decrease in the level of the CTX-II, a marker of cartilage type II collagen degradation.

Thus, in a study of participants with metabolic syndrome who followed a very low-calorie diet (300–600 kcal/day) for nine days, there was an average decrease in body weight of - 4.87 kg, BMI of −2.0, and HOMA-IR index of −3.37 (25). A further investigation of obese women who adhered to a low-calorie diet (∼1200 kcal/day) over a 14-day period revealed that compliance with the diet resulted in an average weight reduction of −2.5 kg and a BMI decrease of −0.9. Additionally, the study observed decreased leptin levels and enhanced insulin sensitivity (26). A study of 12 obese women who were on a low-calorie diet (∼1200 kcal/day) for 2 weeks (one of the study groups) demonstrated an average weight reduction of −2.4 kg, a BMI decrease of −0.8, as well as a significant reduction in fat mass percentage, LDL-C, and insulin (27). Additionally, there is evidence to suggest that a low-calorie diet can influence the progression of osteoarthritis. A study of obese women aged 40 years or older with mild to moderate OA revealed that a low-calorie diet (500 kcal/day less than the estimated intake) for a period of two months led to a significant reduction in body weight, with an average loss of −4.02 kg, as well as a notable decrease in pain levels as measured by VAS. In addition, an anti-inflammatory diet has been shown to be superior to a standard low-calorie diet in its effectiveness in OA (28). A study of 192 obese patients demonstrated that a weight loss program (8 weeks of 415–810 kcal/day and 8 weeks of ∼1200 kcal/day) resulted in a decrease in serum cartilage oligomeric matrix protein (sCOMP), but not in a decrease in CTX-II levels (29).

In this study, we also examined the benefits of adding resveratrol to a low-calorie diet. Overall, we found no benefit of resveratrol on anthropometric parameters or biochemical parameters, including carbohydrate and lipid metabolism, compared to the control group after 10 days of treatment. A significant reduction in hsCRP, an inflammatory marker, was observed. Pain scores decreased in both groups, with the most significant improvement demonstrated in the control group. Changes in the WOMAC score also did not differ between groups. However, resveratrol supplementation did show a more pronounced reduction in urinary CTX-II levels compared to the control group.

A study of overweight and obese participants found that taking resveratrol for four weeks did not improve anthropometric measures (weight and BMI) or blood lipid parameters (30). Another study showed that 30 days of resveratrol supplementation did not result in weight loss. However, it did improve certain biochemical markers, including triglyceride levels and the HOMA index (31). In addition, the results of a study conducted on 24 obese but otherwise healthy men indicated that the administration of a supplement containing 500 mg/day of resveratrol over a period of four weeks did not result in improvements in body composition or biochemical markers (32). Concurrently, an umbrella review of interventional meta-analyses demonstrated minor enhancements in anthropometric parameters (weight, BMI, WC) when resveratrol was administered at a dose exceeding 400 mg/day and for a duration extended beyond 12 weeks (33).

Evidence has been obtained regarding the effect of resveratrol on the progression of OA. Specifically, the administration of 500 mg/day of resveratrol supplements has been shown to result in a significant improvement in pain scores, as measured by the VAS and KOOS, after a 30-day period (34). Additionally, in another study, taking a combination of resveratrol and meloxicam demonstrated benefits in improving pain, functions, and associated symptoms compared with meloxicam alone (35). The lack of concordance between CTX-II reduction and WOMAC scores suggests that biochemical markers of cartilage turnover respond earlier than clinical symptoms in short-term studies. Thus, the study showed that the level of CTX-II is not an independent predictor of some indicators of functional assessment of joint condition, such as WOMAC (36).

We also found an association between increased HDL-C levels and decreased CTX-II. HDL is essential for modulating the development and activation of osteoblasts and osteoclasts, which in turn affects bone integrity (37). A recent study of 4,782 patients hospitalized with osteoporotic fractures also showed this association (38).

It is noteworthy that all study participants were hospitalized during the study period, which contributed to high adherence to the low-calorie diet and enabled significant results to be achieved in a short period of time.

Limitations of this study include the relatively short observation period, which does not allow for objective conclusions to be drawn about the long-term benefits of resveratrol supplementation, and the small sample size. Physical activity during inpatient treatment may have influenced weight loss, pain, inflammation, and metabolic outcomes, but it was not objectively monitored. The cohort included only Caucasian postmenopausal women with obesity and knee osteoarthritis, so the findings may not apply to men, younger patients, other ethnic groups, or non-obese patients.

A strength of this study is the inpatient setting, which ensured close monitoring of participants and high adherence to both the low-calorie diet and resveratrol supplementation.

Future studies should include larger and more diverse populations and account for sex, age, menopausal status, obesity severity, metabolic phenotype, and baseline osteoarthritis stage (39). In addition, future trials should incorporate broader biomarker panels, including markers of cartilage degradation and synthesis, synovial inflammation, bone turnover, oxidative stress, adipokine imbalance, and systemic low-grade inflammation (40). The integration of MRI-based structural outcomes, metabolomic profiling, gut microbiome analysis, and nutrigenetic approaches may help identify patient subgroups most likely to benefit from resveratrol as an adjunct to dietary intervention (41,42).

Overall, these findings suggest that resveratrol may have a limited short-term effect on general metabolic outcomes but may promote favorable changes in inflammation and cartilage degradation markers. Further research is needed to clarify whether these biomarker changes translate into sustained clinical benefits and slower osteoarthritis progression.

## Conclusion

A 10-day low-calorie diet was associated with improvements in anthropometric, metabolic, inflammatory, and osteoarthritis-related parameters in postmenopausal women with obesity and knee osteoarthritis. The addition of resveratrol did not provide clear additional benefits for most clinical and metabolic outcomes, but was associated with greater reductions in hsCRP and urinary CTX-II. Further randomized, placebo-controlled studies with longer follow-up are needed to confirm these findings and clarify the clinical relevance of resveratrol as an adjunct to dietary intervention.

## Funding

The author(s) declared that financial support was received for this work and/or its publication. This publication/research was funded by the Russian Science Foundation, grant number 22-15-00252-P.

## Author contributions CRediT

**Georgy Leonov**: Conceptualization, Formal analysis, Software, Visualization, Writing – original draft; **Anastasia Malvina**: Conceptualization, Investigation, Methodology, Writing – review & editing; **Svetlana Kosyura**: Methodology, Investigation, Data curation, Writing – review & editing; **Elena Livantsova**: Data curation, Writing – review & editing; **Yurgita Varaeva**: Validation, Writing – review & editing; **Antonina Starodubova**: Conceptualization, Funding acquisition, Project administration, Resources, Supervision, Writing – review & editing.

## Acknowledgments

The authors acknowledge the BioRender team for providing the artwork creation online service (BioRender.com).

## Data availability statement

The datasets presented in this manuscript are available from the corresponding author(s) upon reasonable request.

## Conflict of interest

No potential conflict of interest was reported by the author(s).

## Notes

### Competing Interest Statement

The authors have declared no competing interest.

### Clinical Trial

NCT07634302

### Author Declarations

Ethics Committee of the Federal Research Centre of Nutrition, Biotechnology and Food Safety gave ethical approval for this work (protocol No. N1/2017, dated February 3, 2017).

